# The comparative superiority of IgM-IgG antibody test to real-time reverse transcriptase PCR detection for SARS-CoV-2 infection diagnosis

**DOI:** 10.1101/2020.03.28.20045765

**Authors:** Rui Liu, Xinghui Liu, Huan Han, Muhammad Adnan Shereen, Zhili Niu, Dong Li, Fang Liu, Kailang Wu, Zhen Luo, Chengliang Zhu

**Author notes:** **Corresponding authors** Chengliang Zhu, MD, Department of Clinical Laboratory, Renmin Hospital of Wuhan University, 238 Jiefang Road, Wuhan 430060, P. R. China. E-mail address, Zhen Luo, PhD, Guangdong Provincial Key Laboratory of Virology, Institute of Medical Microbiology, Jinan University, 601 Huangpu Avenue, Guangzhou 510632, P. R. China. Rui Liu and Xinghui Liu contributed equally to this work.

## Abstract

**Background:** As the increasing number of Corona Virus Disease 2019 (COVID-19) patients caused by the severe acute respiratory coronavirus 2 (SARS-CoV-2), which caused an outbreak initiated from Wuhan, China in December, 2019, the clinical features and treatment of COVID-19 patients have been understood. However, it is urgent to need the rapid and accurate detection for SARS-CoV-2 infection diagnosis. We aimed to evaluate the antibodies-based and nucleic acid-based tests (NAT) for SARS-CoV-2-infected patients.

**Method:** We retrospectively and observationally studied 133 patients diagnosed with SARS-CoV-2 and admitted in Renmin Hospital of Wuhan University, China, from Feb 17 to Mar 1, 2020. Demographic data, symptoms, clinical examination, laboratory tests, and clinical outcomes were collected. Data were compared between IgM-IgG antibody test and real-time RT-PCR detection for COVID-19 patients.

**Results:** Of 133 patients with SARS-CoV-2 infection, there were 44 moderate cases, 52 severe cases, and 37 critical cases with no significant difference of gender and age among three subgroups. Overall, the positive ratio in IgM antibody test was higher than in RT-PCR detection. In RT-PCR detection, the positive ratio was 65.91%, 71.15%, and 67.57% in moderate, severe, and critical cases, respectively. Whereas, the positive ratio of IgM/IgG antibody detection in patients was 79.55%/93.18%, 82.69%/100%, and 72.97%/97.30% in moderate, severe, and critical cases, respectively. Moreover, the concentrations of antibodies were also measured in three subgroups.

**Conclusion:** The IgM-IgG antibodies-based test exhibited a comparative superiority to the NAT for COVID-19 diagnosis, which provides an effective complement to the false negative results from NAT for SARS-CoV-2 infection diagnosis.

## Introduction

The novel coronavirus (SARS-CoV-2) rapidly spread all over the globe and caused coronavirus disease (COVID-19) in infected persons after its initial emergence in Wuhan, China, in December 2019. SARS-CoV-2 has infected 76,819 people out of which 12,077 were critical, 2251 died (2.9% fatality rate) and 18,878 clinically recovered during the first 50 days of the outbreak (1, 2). To mitigate the risk of spread it is necessary to investigate and develop effective treatment and diagnostic options. The signs and symptoms of SARS-CoV-2 infection are not specific, most are associated with respiratory complications such as cough dyspnea, and viral pneumonia, but the mortality of critically ill patients with SARS-CoV-2 pneumonia is also considerable (3, 4). Therefore, specific COVID-19 diagnostic tests are required to confirm suspected cases. Besides diagnostic techniques, appropriate samples or specimens for the detection of the viral genome are also of high concern (5, 6).

Previous studies on COVID-19 pneumonia have largely focused on clinical characteristics and epidemiology (7, 8). However, very limited details are available related to effective diagnostic strategies. In the current situation, the specificity and sensitivity of the tests are not widely known, therefore, testing of multiple specimen types is recommended (9, 10). The most widely used tests in the current situation are based on nucleic acid detection and antibodies detection. Although the viral nucleic acid RT-PCR test has become the standard method for SARS-CoV-2 infection diagnosis, high false negative rates were reported (11). Upon coronavirus infection, IgM antibodies are produced as an early immune response after infection in the body, which may indicate current infection or new infection. IgG antibodies are the main antibodies produced as an immune response, indicating that the disease has entered a recovery period or that there is a prior infection (12, 13). Therefore, combined tests of immunoglobulin M (IgM) and immunoglobulin G (IgG) antibodies can not only provide early diagnosis of infectious diseases but also help to evaluate the stage of infection in the body (11).

To further facilitate the efforts of clinical staff in testing, we compared the sensitivity and effectiveness of the currently available tests. We reported that serum antibodies-based testing was more sensitive and efficient to be used as a diagnostic option as compared to reverse transcriptase PCR, which exhibited comparatively superior to the nucleic acid-based test for patients in different stages of SARS-CoV-2 infection. Our findings suggested that IgM-IgG antibody test provides an effective complement to the false negative results from nucleic acid test for SARS-CoV-2 infection diagnosis.

## Materials and methods

### Patients

A total number of 133 patients diagnosed with SARS-CoV-2 in Renmin Hospital (Wuhan University, China) from Feb 17 to Mar 1, 2020, were included as the case group. All patients were diagnosed according to the “pneumonia diagnosis protocol for novel coronavirus infection (trial version 5)”, subjected to the tests including clinical examination, Computed Tomography (CT) and real-time reverse-transcription polymerase chain reaction (RT-PCR) for SARS-CoV-2. The SARS-CoV-2 group was divided into three additional subgroups according to new pneumonia diagnosis and treatment of COVID-19 (trial version fifth). The information of three subgroups was divided as 44 moderate cases (22 males and 22 females, median age was 67.5 [64∼71.75]), 52 severe cases (28 males and 24 females, median age was 68 [61.25∼74]), and 37 critical cases (20 males and 17 females, median age was 70 [60∼76.5]). There is no significant difference of sex and age among three subgroups.

### Data collection

Data on biochemical parameters were obtained from all 133 confirmed SARS-CoV-2 infection patients, which was confirmed by a broad series of investigations including clinical examination, laboratory tests, chest x-rays and two independent real-time reverse-transcription polymerase chain reaction (rRT-PCR) for SARS-CoV-2, with SARS-CoV-2 ORF1ab/N qPCR detection kit (GeneoDx Biotech, Shanghai, China), as well as using a SARS-CoV-2 antibody detection kit (YHLO Biotech, Shenzhen, China). Clinical and laboratory information was collected during routine clinical work, and the study was approved by the Ethics Committee and Institutional Review Board of the Renmin Hospital of Wuhan University (certificate no. WDRY2020-K066).

### Statistical analysis

SPSS software version 25.0 was used for statistical analysis. All quantitative data in non-normal or unknown distribution were expressed as median and interquartile range. Wilcoxon rank sum test was used to analyze differences among groups for the measurement data that did not meet the normal distribution. The Chi-square (χ^2^) test was used for the difference between groups of enumeration data. In all tests, *P*<0.05 was defined as statistically significant

## Results

### The value of IgM antibody and RT-PCR detection for SARS-CoV-2 infection diagnosis

The positive ratio was 78.95% (105/133) in IgM antibody test, while 68.42% (91/133) in RT-PCR detection for SARS-CoV-2 infection, respectively, suggesting a higher detection sensibility in IgM antibody test than in RT-PCR test (Table 1). However, there were still false positive and false negative results in two tests. Overall, the sensitivity was higher in case of antibody-based test, while lower sensitivity in case of RT-PCR.

**Table 1.** The comparation of IgM antibody and RT-PCR detection for SARS-CoV-2 infection diagnosis.

| IgM | SARS-CoV-2 RNA |  | Sample Quantity |
| --- | --- | --- | --- |
|  | + | - |  |
| + | 74 | 31 | 105 |
| - | 17 | 11 | 28 |
| Total | 91 | 42 | 133 |

Note: + stands for positive, while – stands for negative.

### The value of RT-PCR detection for viral RNA in COVID-19 patients in different stages

Considering the severity of COVID-19 patients from the critical care resources in hospitals (3), for further analysis in details, the COVID-19 patients was divided into three additional subgroups as 44 moderate cases (22 males and 22 females, median age was 67.5 [64∼71.75]), 52 severe cases (28 males and 24 females, median age was 68 [61.25∼74]), and 37 critical cases (20 males and 17 females, median age was 70 [60∼76.5]) with no significant difference of sex and age among three subgroups (*P*>0.05).

Then, we desired to figure out the RT-PCR detection for viral RNA in three subgroups of COVID-19 patients. In RT-PCR detection for viral RNA in patients infected with SARS-CoV-2, the positive ratio was 65.91% in moderate cases, 71.15% in severe cases and 67.57% in critical cases, respectively (Table 2). Of note, we didn’t observe significant differences in positive ratio among three subgroups of COVID-19 patients.

**Table 2.** The RT-PCR detection for viral RNA in patients infected with SARS-CoV-2.

| SARS-CoV-2<br>RNA | Moderate<br>(n=44) | | Severe<br>(n=52) | | Critical<br>(n=37) | | $\chi^2$ | *P* value |
| --- | --- | --- | --- | --- | --- | --- | --- | --- |
|  | No. (+) | Ratio (+) | No. (+) | Ratio (+) | No. (+) | Ratio (+) |  |  |
| NP | 29 | 65.91% | 38 | 73.08% | 25 | 67.57% | 0.636 | 0.728 |
| ORF1ab | 33 | 75.00% | 42 | 80.77% | 27 | 72.97% | 0.84 | 0.657 |
| NP &<br>ORF1ab | 29 | 65.91% | 37 | 71.15% | 25 | 67.57% | 0.321 | 0.852 |

Note: No., number; Ratio (+), positive ratio.

### The value of IgM-IgG antibody detection for COVID-19 patients in different stages

To further examine the test based on antibody in three subgroups of COVID-19 patients, moderate, severe and critical cases. In IgM antibody detection in patients infected with SARS-CoV-2, the positive ratio was 79.55% in moderate cases, 82.69% in severe cases and 72.97% in critical cases, respectively. Similarly, the positive ratio from IgG antibody test was 93.18% in moderate cases, 100.00% in severe cases and 97.30% in critical cases, respectively (Table 3). We observed the positive ratio was still higher in case of antibody-based test, while lower in case of RT-PCR for the diagnosis of three subgroups of patients.

**Table 3.**
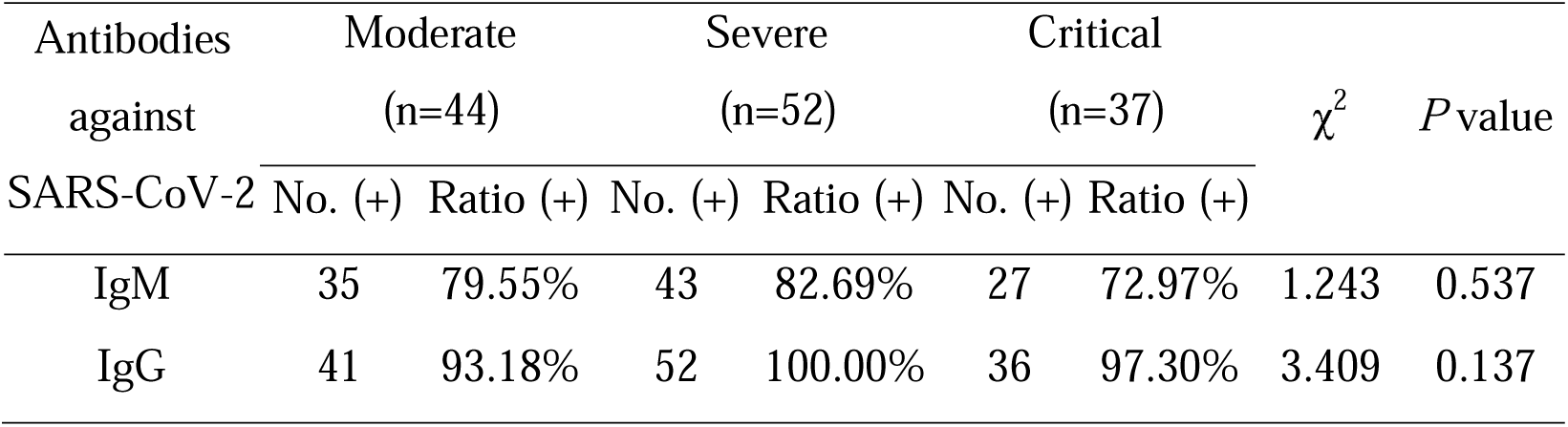
The IgM-IgG antibody detection for patients infected with SARS-CoV-2.

| Antibodies | Moderate | | Severe | | Critical | | $\chi^2$ | *P* value |
| --- | --- | --- | --- | --- | --- | --- | --- | --- |
| against | (n=44) |  | (n=52) |  | (n=37) |  |  |  |
| SARS-CoV-2 | No. (+) | Ratio (+) | No. (+) | Ratio (+) | No. (+) | Ratio (+) |  |  |
| IgM | 35 | 79.55% | 43 | 82.69% | 27 | 72.97% | 1.243 | 0.537 |
| IgG | 41 | 93.18% | 52 | 100.00% | 36 | 97.30% | 3.409 | 0.137 |

### The concentrations of IgM-IgG antibody detection for COVID-19 patients in different stages

Finally, the concentrations of IgM and IgG antibodies in serological test for COVID-19 patients in different stages were measured. The concentration of IgM in patients was 29.19 AU/ml [17.04∼61.02] in moderate cases, 40.76 AU/ml [13.56∼90.13] in severe cases and 23.25 AU/ml [8.67∼104.5] in critical cases, respectively. Meanwhile, the concentration of IgG in patients was 147.73 AU/ml [89.53∼171.6] in moderate cases, 148.63 AU/ml [130.95∼167.7] in severe cases and 140.4 AU/ml [93.79∼162.8] in critical cases, respectively (Table 4). Collectively, there were no significant differences in antibodies concentrations among three subgroups of COVID-19 patients, but the test results still revealed as a considerable diagnosis for COVID-19 progression.

**Table 4.** The comparation of concentrations of IgG and IgM antibodies (AU/ml) in patients infected with SARS-CoV-2.

| Antibodies<br>against | Moderate | Severe | Critical | *P* value |
| --- | --- | --- | --- | --- |
| SARS-CoV-2 | n=44 | n=52 | n=37 |  |
| IgM | 29.19<br>(17.04 ~ 61.02) | 40.76<br>(13.56 ~ 90.13) | 23.25<br>(8.67 ~ 104.5) | 0.446 |
| IgG | 147.73<br>(89.53 ~ 171.6) | 148.63<br>(130.95 ~ 167.7) | 140.4<br>(93.79 ~ 162.8) | 0.182 |

Note: The concentration unit of antibodies in serum samples is AU/ml. The value of AU/ml >10 is considered as positive reaction.

## Discussion

The outbreak of pneumonia caused by SARS-CoV-2 spreads rapidly, posing a serious threat to the lives and health of the people. SARS-CoV-2 belongs to the coronavirus beta genus, with a linear single-stranded positive-chain RNA, the seventh coronavirus known to infect humans after SARS (2002) and MERS (2012) (14). There are various assays developed to detect different regions of the SARS-CoV-2 genome using RT-PCR (9, 15). In the present study, we applied both antibody and nucleic acid based diagnostic strategies on suspected patients with moderate to critical symptoms for COVID-19. Total 133 patients were tested, where 68.42% (91/133) were positive in case of RT-PCR and 78.95% (105/133) in case of antibody test. It was observed that antibody testing was rapid and had significantly higher efficiency and sensitivity.

Recently, chest CT scans were applied for the rapid detection of SARS-CoV-2 induced COVID-19 (10, 16). The chest X-ray or chest CT provides more information, but these are not conclusive as not all the patients with COVID-19 develop pneumonia, and many other things can cause pneumonia (17, 18). Therefore, a more effective strategy such that testing antibodies or RNA is important. The conventional serologic assays and CRISPR-nCoV based detection is also a novel approach for the detection of SARS-CoV-2 (11, 19, 20). As SARS-CoV-2 is a new infectious disease and the immunological testing reagents have recently been developed (11). Although the antibodies generated after a period of the onset of infection, their detections were found more promising in the current situation.

The IgM and IgG testing in combination are of great value for improving the clinical sensitivity of early COVID-19 diagnosis. Certainly, it was been confirmed that the detection sensibility was higher in IgG-IgM combined antibody test than in individual IgG or IgM antibody test (11). In general, the coronavirus stimulates the immune response and IgM antibodies are produced firstly and then quickly decline until disappear, while on the other hand, IgG antibodies are usually produced after IgM and continue to rise and remain high in the body for long periods of time (12, 13). For treatment monitoring and status of the disease, the decrease or even disappearance of the concentration of IgM and the increase in the concentration of IgG indicates the severity of the patient and the immunity to the pathogenicity of SARS-CoV-2. Therefore, further investigations should be made on a broad range and mainly focus on the antibody’s response pattern and severity status of the patient on the bases of antibodies production.

In conclusion, the higher sensitivity for IgM/IgG antibody-based testing may be associated with its concentration level. The higher level of confirmation of infection in severe cases, the higher sensitivity, and lesser false negative results indicate that diagnostic testing based on IgG has the potential to be accepted widely. Considering the significance of this ongoing COVID-19 epidemic and risk of pandemics, we believe that our findings are important in terms of providing the promising diagnostic options based on age and sex groups, as well as the severity of symptoms. We further recommend IgM-IgG antibody test provides an effective complement to the false negative results from nucleic acid test for SARS-CoV-2 infection diagnosis.

## Data Availability

All data in the study appear in the submitted article.

## Acknowledgement

This work was supported by the National Natural Science Foundation of China (81672079 to CZ and 31800147 to ZL), the Open Research Fund Program of the State Key Laboratory of Virology of China (2019KF001 to ZL), the Outstanding Leaders Training Program of Pudong Health Bureau of Shanghai (PWR12018-05 to XL), and the Key Disciplines Group Construction Project of Pudong Health Bureau of Shanghai (PWZxq2017-15 to XL).

## Author contributions

All the authors have accepted responsibility for the entire content of this submitted manuscript and approved submission.

